# Effects of Diazepam on Hippocampal Blood Flow in People at Clinical High Risk for Psychosis

**DOI:** 10.1101/2024.01.03.23299658

**Authors:** Nicholas R. Livingston, Amanda Kiemes, Gabriel A. Devenyi, Samuel Knight, Paulina B. Lukow, Luke A. Jelen, Thomas Reilly, Aikaterini Dima, Maria Antonietta Nettis, Cecilia Casetta, Tyler Agyekum, Fernando Zelaya, Thomas Spencer, Andrea De Micheli, Paolo Fusar-Poli, Anthony A. Grace, Steve C.R. Williams, Philip McGuire, Alice Egerton, M. Mallar Chakravarty, Gemma Modinos

## Abstract

**Background:** Elevated hippocampal perfusion has been observed in people at clinical high-risk for psychosis (CHR-P). Preclinical evidence suggests that hippocampal hyperactivity is central to the pathophysiology of psychosis, and that prophylactic treatment with diazepam during adolescence can prevent the development of psychosis-relevant phenotypes. Here we examined whether diazepam normalises hippocampal perfusion in CHR-P individuals.

**Methods:** Using a randomised, double-blind, placebo-controlled, crossover design, 24 CHR-P individuals were assessed with MRI on two occasions, once following a single oral dose of diazepam (5 mg) and once following placebo. Regional cerebral blood flow (rCBF) was measured using 3D pseudo-continuous arterial spin labelling and sampled in native space using participant-specific hippocampus/subfield masks (CA1, subiculum, CA4/dentate gyrus). Twenty-one healthy controls (HC) were scanned using the same acquisition sequence, but without administration of diazepam or placebo. Mixed-design ANCOVAs and linear mixed-effects models were used to examine the effects of group (CHR-P placebo/diazepam vs. HC) and condition (CHR-P diazepam vs. placebo) on rCBF in the hippocampus as a whole and by subfield.

**Results:** Under the placebo condition, CHR-P individuals (mean[±SD] age: 24.1[±4.8] years, 15F) showed significantly elevated rCBF compared to HC (mean[±SD] age: 26.5[±5.1] years, 11F) in the hippocampus (*F*(1,36)=8.2, *p*_FDR_=0.004) and across its subfields (all *p*_FDR_<0.001). Following diazepam, rCBF in the hippocampus (and subfields, all *p*_FDR_<0.001) was significantly reduced (*t*(69)=-5.1, *p*_FDR_<0.001) and normalised to HC levels (*F*(1,41)=0.3, *p*_FDR_=0.225).

**Conclusions:** Diazepam can normalise hippocampal hyperperfusion in CHR-P individuals, consistent with evidence implicating medial temporal GABAergic dysfunction in the pathophysiology of psychosis.

## Introduction

Transformations in our understanding of the nature, aetiology, and early course of psychotic disorders drove a marked re-orientation of mental health services toward early intervention in individuals at clinical high-risk for psychosis (CHR-P), raising the prospect that prevention of psychosis onset may be a realistic goal^1,2^. Despite this progress, current treatments have a minimal influence on transition to psychosis^3,4^, and there is no robust evidence to favour any single preventive intervention over another^5,6^. A better understanding of the neurobiology underlying the CHR-P phenotype is critical for the much-needed development of interventions to prevent psychosis onset.

Postmortem, preclinical, and clinical neuroimaging evidence suggests that hippocampal abnormalities are central to the pathophysiology of psychosis^7,8^, thus representing a potential therapeutic target. Hippocampal dysfunction in psychosis is proposed to arise from a disruption in neural excitatory/inhibitory balance driven by GABAergic inhibitory interneuron dysfunction^9–13^. This would render the hippocampus dysrhythmic and hyperactive^14^, and excessive glutamatergic output^15–18^ from the hippocampus to the striatum, amygdala, and prefrontal cortex may underlie the development of positive, negative, and cognitive symptoms, respectively^19^. Preclinical findings in the well-validated methylazoxymethanol acetate (MAM) rodent model of neurodevelopmental disruption^20–22^ indicate that a loss of hippocampal parvalbumin-expressing (PV+) inhibitory interneurons^23^ leads to hippocampal hyperactivity^24,25^, as measured with electrophysiology. Rodent models have also shown that selective reduction in PV mRNA expression^26^ or knock-out of PV+ interneuron expression^24,25^ are each sufficient to induce hippocampal hyperactivity. In MAM-treated rats, hippocampal hyperactivity leads to striatal hyperdopaminergia^21^, which is a core neurobiological feature of positive symptoms in schizophrenia^19^. This hippocampal hyperactivity can in turn be normalised by hippocampal chemical inactivation^22,27^, or by pharmacologically facilitating hippocampal GABA signalling through direct infusion of either a nonselective (the benzodiazepine midazolam)^28^ or an α5-subunit selective GABA_A_ receptor positive allosteric modulator (PAM)^28,29^. Furthermore, chronic administration of the benzodiazepine diazepam during the peripubertal period prevented the development of psychosis-relevant features in adulthood^30–32^, highlighting the prophylactic potential of GABA-enhancing compounds for psychosis.

In individuals at CHR-P, hippocampal hyperactivity has been observed *in vivo*^33–37^, which – as a result of neurovascular coupling – is associated with increased regional cerebral blood flow or volume (rCBF or rCBV), revealed using arterial spin labelling (ASL) or gadolinium-contrast magnetic resonance imaging (MRI), respectively. Elevated hippocampal rCBF in CHR-P individuals has been associated with elevated striatal pre-synaptic dopamine synthesis capacity^38^ and elevated medial prefrontal cortex GABA concentrations^39^. Baseline elevated hippocampal rCBF or rCBV was also found to predict higher positive and negative symptoms severity^33^, poor functioning^33,38^, and transition to psychosis^33,34,39^, and to normalise in those individuals who remit from the CHR-P state^35^. Within the hippocampus, hyperactivity is proposed to originate in the CA1 subfield and then extend to the subiculum and beyond^33,34,37^. Hence, hippocampal rCBF may be a biomarker for symptom severity and psychosis onset in CHR-P individuals, which preclinical evidence suggests may be prevented by pharmacological enhancement of hippocampal GABA levels^30–32^.

To determine whether pharmacologically targeting hippocampal dysfunction in CHR-P individuals with a GABA-enhancing drug may prevent or delay the onset of psychosis, proof-of-concept mechanistic evidence is first required to elucidate its effects on hippocampal hyperactivity. While prior positron or xenon emission tomography (PET or XET) research demonstrated global reductions in CBF under an acute non-sedating dose of a benzodiazepine in healthy controls^40–43^, including reductions in temporal lobe rCBF^40^, no studies have investigated the hippocampus specifically. Furthermore, no studies have been conducted in CHR-P individuals, in whom the effects of a benzodiazepine challenge on hippocampal rCBF may differ from that in healthy controls given the proposed GABAergic dysfunction in this population^14^. Hence, the present study compared the acute effects of a single dose of diazepam vs. placebo on ASL-derived rCBF in the hippocampus and its subfields in 24 CHR-P individuals. To determine baseline alterations in the CHR-P group, rCBF measures from CHR-P individuals under placebo were compared to those from 21 healthy controls (HC). We hypothesised that compared to HC, CHR-P individuals under placebo would show increased rCBF in the hippocampus and its subfields, which would be most apparent in the CA1 subregion^33,34^. Secondly, consistent with the preclinical evidence^29^, we hypothesised that a single dose of diazepam would reduce hippocampal rCBF in CHR-P individuals compared to placebo, and that this would be observed across all subfields due to similar levels of GABA_A_ receptor expression^44,45^. For completeness, supplementary analyses examined broader effects of diazepam on voxel-wise grey matter (GM) rCBF, as well as associations between baseline levels of symptoms/functioning and diazepam-induced hippocampal rCBF changes in CHR-P individuals.

## Methods and Materials

### Participants

#### CHR-P individuals

Twenty-four CHR-P individuals, aged 18-32, were recruited from OASIS (Outreach and Support in South London), an early-intervention service within the South London and Maudsley NHS Foundation Trust, UK^46^. CHR-P criteria was assessed using the Comprehensive Assessment of At-Risk Mental State (CAARMS)^47^. All individuals were required to be experiencing current attenuated psychotic symptoms, defined as having a severity and frequency score of 3 or above for an ongoing symptom on either P1 (unusual thought content), P2 (non-bizarre ideas), P3 (perceptual abnormalities), or P4 (disorganised speech) of the CAARMS. Exclusion criteria included previous/current exposure to antipsychotics, current exposure to psychotropic medications with GABAergic or glutamatergic action (except for antidepressants), pregnancy/breastfeeding, IQ<70, and any contraindication to MRI scanning.

#### HC individuals

In order to validate hippocampal hyperactivity in this sample of CHR-P individuals, we utilised data from 21 HC from a previous study^48^, scanned on the same MRI scanner with the same scanning sequences and acquisition parameters as the CHR-P sample (below). HC data were reanalysed with the same preprocessing steps as the CHR-P data. Details on the HC dataset recruitment and inclusion/exclusion criteria are described in the original publication^48^.

### Study design and procedure

This study had full ethical approval from the National Health Service UK Research Ethics Committee and was carried out at King’s College London. All participants provided written informed consent. Using a randomised, double-blind, placebo-controlled, crossover design, the 24 CHR-P participants underwent two MRI sessions, once under an acute oral dose of diazepam (5 mg; generic) and once under oral placebo (50 mg ascorbic acid; Crescent Pharma Ltd, Hampshire, UK), with a minimum 3-week washout period between visits. Prior to the first scanning visit and randomisation, during an assessment visit demographic information, basic medical history, and clinical/cognitive assessments were collected, including the CAARMS^47^, Global Functioning Social and Role scales^49^, Hamilton Anxiety and Depression scales^50,51^, Wechsler Adult Intelligence Scale III^52^, and the Trail Making Test A & B^53^. At each scanning visit, the capsule (with diazepam or placebo) was administered 60 minutes prior to MRI scanning, so that the MRI session coincided with peak diazepam plasma levels^54^. Full details of the clinical/cognitive assessments and scanning visit procedures (including randomisation and maintenance of blinding) can be found in the Supplementary Methods.

### MRI acquisition

MRI scanning for all participants was conducted at the Centre for Neuroimaging Sciences, King’s College London, using a General Electric MR750 3.0T MR scanner with an 8-channel head coil. rCBF was measured using a 3D pseudo-continuous ASL sequence (pcASL; TE=11.088 ms, TR=4,923 ms, FoV=240) as used in previous CHR-P studies^35,36,48^. Total scan time was 6 minutes and 8 seconds. Further details of the pcASL sequence can be found in the Supplementary Methods. For image registration and generation of hippocampal/subfield masks, a T1-weighted SPGR (TE=3.02 ms, TR=7.31 ms, TI=400 ms, flip angle=11°, matrix=256×256, FoV=270, in-plane resolution=1.05×1.05 mm^2^, slice thickness=1.2 mm, 196 slices) and T2-weighted FRFSE (TE=64.9 ms, TR=4380 ms, flip angle=111°, matrix=320×256, FoV=240, in-plane resolution=1.25×0.94 mm^2^, slice thickness=2 mm, 72 slices) image was acquired.

### Image processing

#### Generation of hippocampal/subfield and total GM masks

Structural scans were preprocessed, including denoising, and correcting bias field inhomogeneities using the N3 algorithm^55^. Hippocampus/subfield masks (whole hippocampus, subiculum, CA1, and CA4/DG) were then generated for each participant from their preprocessed structural T1 SPGR scan by using the MAGeT Brain (multiple automatically generated templates of different brains) toolbox^56–58^ (Figure 1). This toolbox has been validated to generate accurate hippocampus and subfield segmentations in Alzheimer’s disease and psychosis cohorts, with greater accuracy than other available toolboxes including Freesurfer 7 and FSL FIRST^57^. This is largely due to an intermediate template step which allows incorporating the neuroanatomical variability of the dataset into the segmentation of each participant, reducing registration and resampling errors, thereby yielding more accurate results. Hippocampal subfields CA2/3 were not included due to the limitations in reliably sampling these smaller regions within the spatial resolution and low signal-to-noise ratio constraints of ASL^59^. Total GM masks were made through binarising GM segmentations.

**Figure 1.**
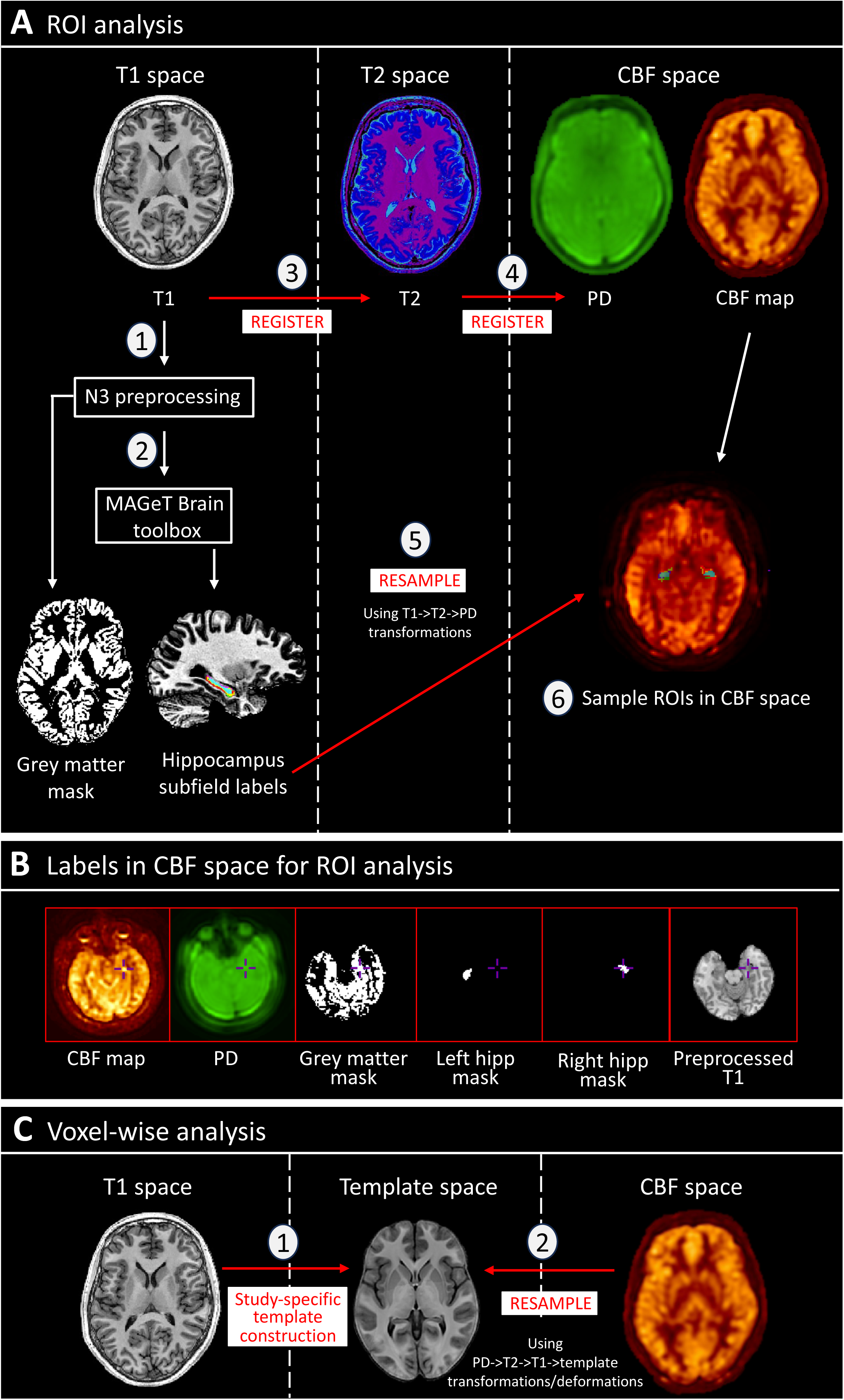
ASL preprocessing and analysis pipeline. (A) Diagram showing pipeline for region-of-interest analysis. T1 images were preprocessed (1) and run through the MAGeT Brain toolbox (2), generating masks for grey matter, whole hippocampus, and hippocampal subfields. Using T1->T2->PD transformations calculated from registration (3 & 4), these masks were resampled (5) to CBF space (single resampling step) to allow for sampling of rCBF in native space (6). **(B)** Demonstration of optimum registration between CBF map, masks, and T1 images. **(C)** Schematic showing steps for voxel-wise analysis for CHR-P placebo vs. diazepam. A study-specific template was generated (1) from participant-averages (averaged structural scans from both drug conditions), and the CBF maps were resampled (2) to common space (single resampling step calculated from PD->T2->T1->template transformations/deformations). hipp: hippocampus; PD: proton density; rCBF: regional cerebral blood flow; ROI: region-of-interest.

#### ASL sampling

Masks were registered and resampled to the individuals respective CBF map using ANTs/2.5.0 (https://github.com/ANTsX/ANTs), and the mean rCBF value was extracted per ROI per hemisphere in native space using minc-toolkit-v2/1.9.18 tools (https://bic-mni.github.io/; Figure 1; Supplementary Methods).

#### Region-of-interest (ROI) analysis

All ROI analyses were completed in R 4.2.2 (https://www.r-project.org/). Individual models assessed the effect of group (CHR-P placebo/diazepam vs. HC) or condition (CHR-P diazepam vs. placebo) on rCBF per ROI (total GM, whole hippocampus, CA1, subiculum, CA4/DG; Figure 2C). Each model included rCBF values per hemisphere, on the basis that bilateral sampling of the same region in the same subject reflects a repeated measurement. Therefore, a group/condition*hemisphere term was included to investigate whether the effect of group or condition on rCBF significantly differed within a region based on hemisphere. Significance was set at pFDR<0.05, after correcting for multiple comparisons using the Benjamini-Hochberg method^60^.

**Figure 2.**
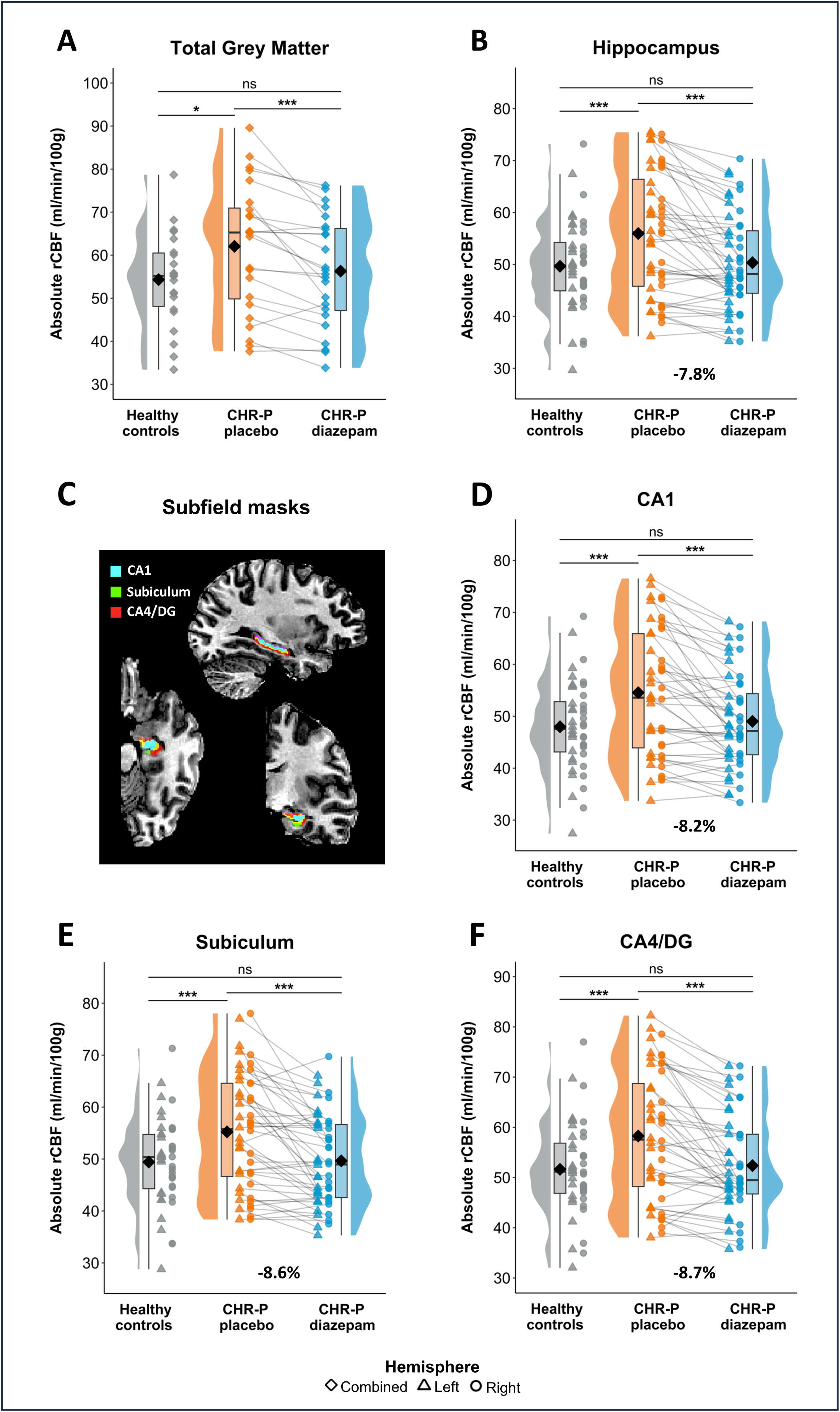
Region-of-interest rCBF findings. Absolute rCBF for HC and CHR-P participants (under placebo and diazepam) for (**A**) total grey matter, (**B**) hippocampus, and (**C**) hippocampus subfields, including (**D**) CA1, (**E**) subiculum, and (**F**) CA4/DG. CHR-P: clinical high-risk for psychosis; DG: dentate gyrus; rCBF: regional cerebral blood flow ns = non-significant; * < 0.05; *** < 0.001.

##### CHR-P placebo vs. HC

Two-way mixed analysis of covariance (ANCOVA; package stats/3.6.2) models assessed baseline rCBF differences in CHR-P individuals compared to HC, covarying for age, sex, and global CBF. Supplementary analyses were performed covarying for demographic characteristics which differed between groups (IQ, ethnicity) or are known to impact rCBF (daily current cigarette use^61^ and ROI grey matter volume^62^).

##### CHR-P diazepam vs. CHR-P placebo

Diazepam-induced changes in rCBF compared to placebo were assessed using linear mixed-effects models (package lme4/1.1-34) with participant ID as a random effect (intercept). Given the within-subject design and widespread expression of GABA_A_ receptors with benzodiazepine binding sites across the brain^44^, global CBF was not included as a covariate. Supplementary analyses investigated potential confounding effects of global CBF, sex, order of scan conditions, and number of days between scans on the results.

##### CHR-P diazepam vs. HC

To examine whether diazepam would normalise hippocampal rCBF in CHR-P individuals to HC levels, two-way mixed ANCOVA models were run covarying for age, sex, and global CBF.

### Exploratory/supplementary analyses

#### Voxel-wise grey matter rCBF analysis

For completeness, we explored the effects of diazepam vs. placebo on voxel-wise GM rCBF. The two-level modelbuild toolkit (github.com/CoBrALab/optimized_ants MultivariateTemplateConstruction) was used to generate a study-specific anatomical template (Figure 1; Supplementary Methods). CBF maps were resampled into common-space and smoothed with a 6 mm FWHM Gaussian kernel. A voxel-wise linear mixed-effects model (R-3.5.1, RMINC-1.5.2.2, lme4 1.1-21) was used to investigate the effect of condition (CHR-P diazepam vs. placebo) on rCBF, with participant ID as random effect and masked using a study-average GM mask. Significance was set at pFDR<0.05. The above procedure was repeated for investigating voxel-wise group differences in rCBF between CHR-P placebo and HC, using the one-level model template build and running a voxel-wise linear model, covarying for global CBF, age, and sex, and masked using a study-average GM mask.

#### Clinical characteristics and hippocampal rCBF change

Supplementary analyses using Pearson’s correlations assessed whether baseline clinical characteristics (positive symptom severity, negative symptom severity, cognitive functioning, social functioning, role functioning, anxiety symptom severity, depression symptom severity) were associated with the mean percent change in bilateral hippocampal rCBF under diazepam vs. placebo (see Supplementary Methods for further details on composition of clinical characteristics scores). Potential confounding effects of global CBF on these results were explored using partial Pearson’s correlations. Outlier detection was performed on significant correlations using Cook’s distance. Significance was set at *p*<0.05, and multiple comparison corrections were not performed as these analyses were exploratory.

## Results

Participant demographic and clinical characteristics are detailed in Table 1. CHR-P individuals had a significantly lower IQ compared to HC and differed in terms of ethnicity. This was driven by an above-average mean IQ^63^ and a higher proportion of white ethnicity in the HC group.

**Table 1.** Demographic and clinical characteristics. CAARMS: comprehensive assessment of at-risk mental states; CHR-P: clinical high-risk for psychosis; HC: healthy control; IQ: intelligent quotient; WAIS: Wechsler adult intelligence scale.

| | CHR-P<br>(n = 24) | HC<br>(n = 21) | *t*/$\chi^2$ | *p* |
| --- | --- | --- | --- | --- |
| Age (years; mean ±SD) | 24.1 ±4.8 | 26.5 ±5.1 | 1.7 | 0.09 |
| Sex (male/female; n) | 9/15 | 10/11 | 0.4 | 0.49 |
| Ethnicity (n) |  |  | 13.9 | **0.007** |
| White | 11 | 16 | - | - |
| Asian | 2 | 6 | - | - |
| Black | 6 | 0 | - | - |
| Mixed or multiple | 4 | 0 | - | - |
| Other | 1 | 0 | - | - |
| IQ (WAIS-III short version<sup>63</sup>; mean ±SD) | 97.6 ±21.6 | 119.3 ±16.7 | 4.6 | **<0.001** |
| Current daily cigarette use, n (%) | 8 (33) | 2 (9) | 3.7 | 0.055 |
| Current alcohol use, n (%) | 18 (75) | 19 (90) | 1.8 | 0.177 |
| Current cannabis use, n (%) | 7 (29) | 4 (19) | 0.6 | 0.431 |
| CAARMS<sup>47</sup> score (mean ±SD) | | | | |
| Positive symptoms | 46.4 ±12.9 | NA | - | - |
| Negative symptoms (n=21) | 29.1 ±24.7 | NA | - | - |
| Total (n=21) | 75.9 ±29.9 | NA | - | - |
| Global functioning score<sup>49</sup> (mean ±SD) | | | | |
| Social | 6.4 ±1.5 | NA | - | - |
| Role | 6.1 ±1.8 | NA | - | - |
| Hamilton scale score (mean ±SD) | | | | |
| Anxiety<sup>50</sup> (n=22) | 17.1 ±8.5 | NA | - | - |
| Depression<sup>51</sup> (n=21) | 13.9 ±6.9 | NA | - | - |
| Current antidepressant medication, n (%) | 9 | NA | - | - |
| Current or prior antipsychotic medication, n (%) | 0 | NA | - | - |
| Current benzodiazepine/hypnotic medication, n (%) | 0 | NA | - | - |

CAARMS: comprehensive assessment of at-risk mental states; CHR-P: clinical high-risk for psychosis; HC: healthy control; IQ: intelligent quotient; WAIS: Weschler adult intelligence scale

### ROI analysis

#### CHR-P placebo vs. HC

##### Global CBF

Mean CBF in total brain GM (ml/100g/min) was significantly higher (*F*(1,42)=5.2, *p*_FDR_=0.014) in CHR-P individuals under placebo (62.1±14.9) compared to HC (54.3±10.9; Figure 2A).

##### Hippocampus and subfield rCBF

After controlling for global CBF, age, and sex, CHR-P participants in the placebo condition had significantly higher rCBF compared to HC (12.8%) in the hippocampus (*F*(1,41)=24.7, *p*_FDR_<0.001), which did not differ between hemispheres (*F*(1,44)=1.6, *p*_FDR_=0.217; Figure 2B). Similar results were found for all subfields: **CA1** (group: *F*(1,41)=25.8, *p*_FDR_<0.001; group*hemisphere: *F*(1,44)=1.1, *p*_FDR_=0.364; Figure 2D), **subiculum** (group: *F*(1,41)=25.7, *p*_FDR_<0.001; group*hemisphere: *F*(1,44)=1.2, *p*_FDR_=0.274; Figure 2E), and **CA4/DG** (group: *F*(1,41)=20.4, *p*_FDR_<0.001; group*hemisphere: *F*(1,44)=3.2, *p*=0.082; Figure 2F). These results did not change when adding in additional covariates (IQ, ethnicity, current daily cigarette use, ROI grey matter volume; Supplementary Table 1).

#### CHR-P diazepam vs. CHR-P placebo

##### Global CBF

In CHR-P participants, mean CBF in total brain grey matter was significantly lower (*t*(23)=-4.3, *p*_FDR_<0.001) under diazepam (56.3±12.7) compared to placebo (62.1±14.9; Figure 2A).

##### Hippocampus and subfield rCBF

Diazepam significantly reduced rCBF in the hippocampus by 7.8% (*t*(69)=-5.1, *p*_FDR_<0.001), which did not differ between hemispheres (*t*(69)=0.9, *p*_FDR_=0.366; Figure 2B). This effect was observed across all subfields: **CA1** 8.2% (condition: *t*(69)=-5.1, *p*_FDR_<0.001; condition*hemisphere: *t*(69)=0.8, *p*_FDR_=0.403; Figure 2D), **subiculum** 8.6% (condition: *t*(69)=-4.9, *p*_FDR_<0.001; condition*hemisphere: *t*(69)=1.1, *p*_FDR_=0.303; Figure 2E), and **CA4/DG** 8.7% (condition: *t*(69)=-4.7, *p*_FDR_<0.001; condition*hemisphere: *t*(69)=0.8, *p*=0.405; Figure 2F). These results did not change after controlling for global CBF, sex, order of scan conditions, or number of days between scans (Supplementary Table 1).

#### CHR-P diazepam vs. HC

##### Global CBF

There was no significant difference (*F*(1,42)=0.5, *p*_FDR_=0.209) in mean total brain GM CBF in CHR-P in the diazepam condition (56.3±12.7) compared to HC (54.3±10.9; Figure 2A).

##### Hippocampus and subfield rCBF

There was no significant difference in rCBF between CHR-P under diazepam compared to HC in the hippocampus (*F*(1,41)=0.4, *p*_FDR_=0.204; Figure 2B), CA1 (*F*(1,41)=0.9, *p*_FDR_=0.153; Figure 2D), subiculum (*F*(1,41)=0.8, *p*_FDR_=0.272; Figure 2E), or CA4/DG (*F*(1,41)=0.263, *p*_FDR_=0.201; Figure 2F).

#### Exploratory/supplementary analyses

##### Voxel-wise grey matter rCBF analysis

###### CHR-P placebo vs. HCS

Several cortical regions showed increases (e.g., inferior/dorsolateral frontal gyrus and temporal pole, p_FDR_<0.05) and decreases (e.g., inferior parietal and middle occipital gyrus, p_FDR_<0.05) in rCBF in the CHR-P placebo condition compared to HC (Supplementary Figure 1; Supplementary Table 2).

###### CHR-P diazepam vs. CHR-P placebo

There was a global pattern of reduced rCBF under the diazepam compared to placebo condition across cortical and subcortical areas (Figure 3; Supplementary Table 3). Peak voxels (all p_FDR_<0.01) were located in temporal (temporal pole, parahippocampal gyrus, hippocampus, amygdala, middle temporal gyrus), parietal (pre/post central gyrus, middle cingulate), frontal (dorsolateral prefrontal cortex (PFC), ventromedial orbitofrontal cortex, insula, superior frontal gyrus), and occipital (lingual gyrus, occipital gyrus, cuneus) regions, cerebellum, and subcortical regions (thalamus, putamen, caudate, and nucleus accumbens).

**Figure 3.**
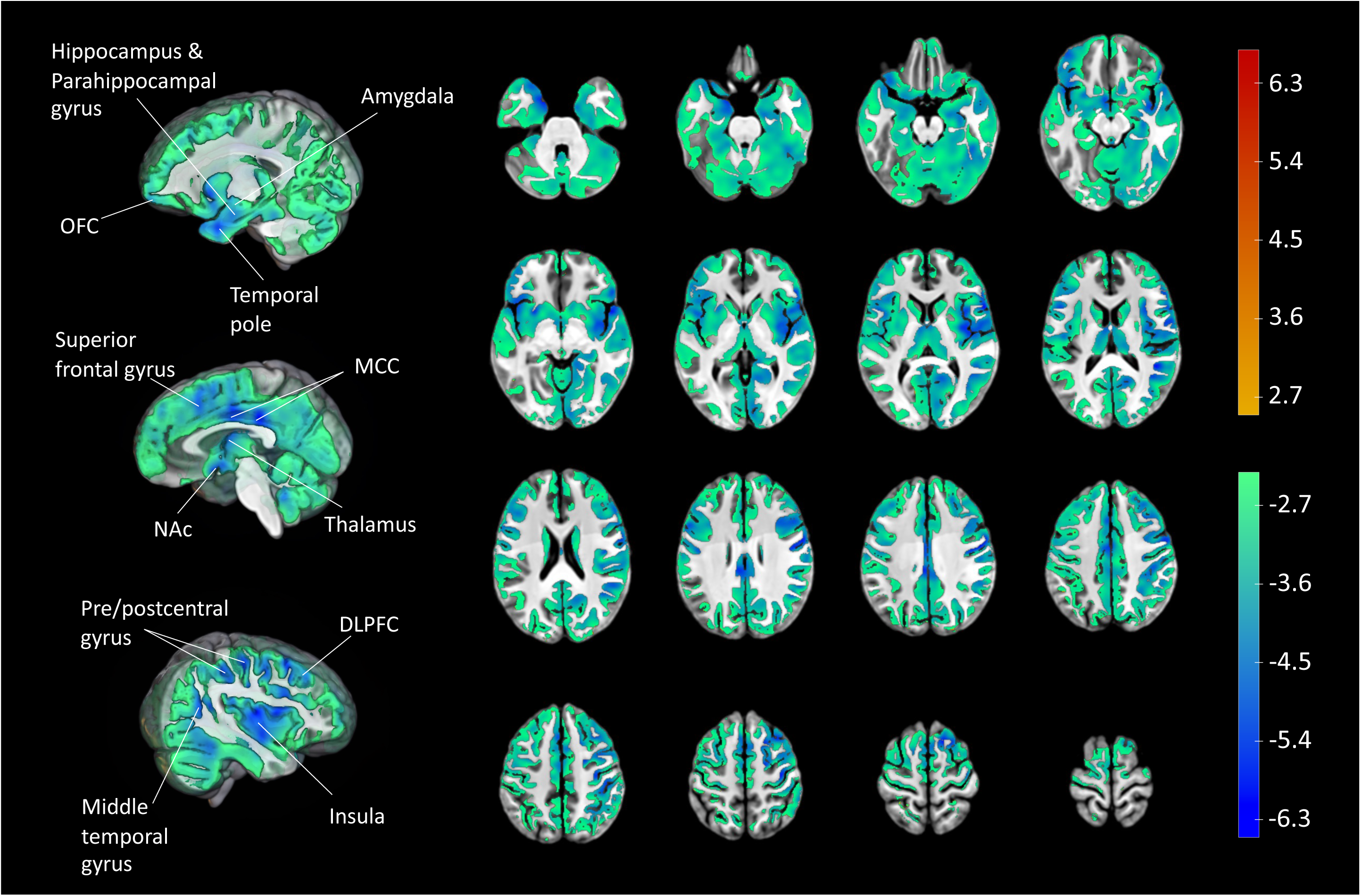
Voxel-wise grey matter rCBF findings. *T*-statistic map of drug condition (diazepam vs. placebo) effect on grey matter rCBF at the whole-brain level in CHR-P individuals from voxel-wise linear mixed effects models, thresholded and displayed at 5% FDR. Peak regions with *t*-statistic > 5 have been labelled. Colour bars denote *t*-statistics which reflect 5% FDR threshold (i.e., ±2.498) and less (i.e., up to ±6.68) for both contrasts (diazepam < placebo in blue/green and placebo < diazepam in yellow/red). N.B. there were no significant voxels at 5% FDR threshold for placebo < diazepam. DLPFC: dorsolateral prefrontal cortex; MCC: middle cingulate cortex; NAc: nucleus accumbens; OFC: orbitofrontal cortex; rCBF: regional cerebral blood flow

#### Clinical characteristics and hippocampal rCBF change

Exploratory analyses revealed significant associations between baseline positive symptoms severity/social functioning and change in hippocampal rCBF, such that higher baseline positive symptom severity (*r*=0.494, *p*=0.014; Figure 4A) and poorer social functioning (*r*=- 0.416, *p*=0.043; Figure 4D) were associated with less reduction in hippocampal rCBF following diazepam vs. placebo. Removal of outliers strengthened these correlations (positive symptoms: *r*=0.598, *p*=0.003; social functioning: *r*=-0.538, *p*=0.008). There were no significant correlations between negative symptoms (*r*=-0.188, *p*=0.415; Figure 4B), cognitive symptoms (*r*=0.114, *p*=0.641; Figure 4C), role functioning (*r*=-0.106, *p*=0.623; Figure 4E), anxiety (*r*=-0.217, *p*=0.333; Figure 4F) or depression symptoms (*r*=-0.155, *p*=0.501; Figure 4G) and diazepam-induced change in whole hippocampal rCBF. Most of these results remained unchanged when adding global CBF as covariate (Supplementary Table 4).

**Figure 4.**
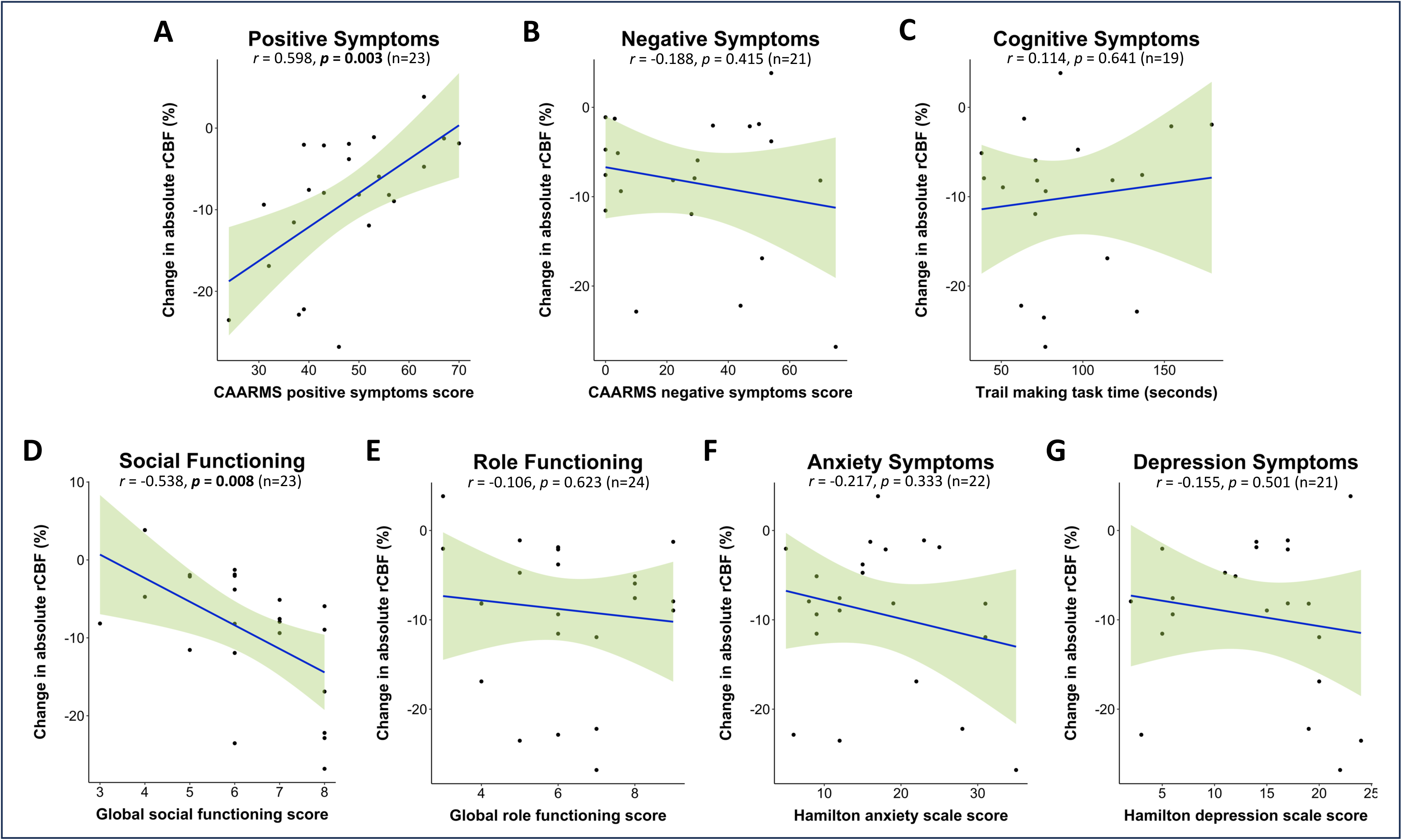
Association between baseline clinical characteristics and diazepam-induced hippocampal rCBF changes. Pearson correlations between change in absolute hippocampal rCBF by diazepam vs. placebo and baseline clinical characteristics (at assessment visit): **(A)** positive symptoms, **(B)** negative symptoms, **(C)** cognitive functioning, **(D)** social functioning, **(E)** role functioning, **(F)** anxiety symptoms, and **(G)** depression symptoms. N.B. for panels D & E a higher score denotes *less* impairment, whilst for all other panels a higher score/time denotes *higher* symptom severity. Shaded light green areas reflect 95% confidence intervals. rCBF: regional cerebral blood flow.

## Discussion

In our sample, hippocampal rCBF was significantly higher in CHR-P individuals under placebo compared to HC. Following diazepam, hippocampal rCBF in CHR-P individuals was significantly reduced and normalised to HC levels. This effect was evident across all hippocampal subfields investigated. Our findings in CHR-P individuals present forward-translation of mechanistic evidence from psychosis-relevant preclinical studies, lending new empirical support to the notion that regulating hippocampal hyperactivity through GABA-enhancing compounds may be a potential therapeutic strategy in early psychosis.

Our finding of elevated hippocampal rCBF in CHR-P (under placebo) compared to HC is consistent with previous rCBF or rCBV studies^33–37^. We also identified a global increase in CBF, as previously reported^35^. Nonetheless, hippocampal rCBF elevation remained significant after controlling for global CBF, suggesting that the hippocampus is particularly hyperactive in CHR-P individuals compared to controls. We hypothesised that elevations in hippocampal rCBF would be most pronounced in the CA1 subfield, based on a recent rCBV study in CHR-P individuals^37^ and current models of psychosis pathophysiology^7^. However, we observed similarly elevated rCBF across all subfields (CA1, subiculum, and CA4/DG). We sampled rCBF across the whole length of the hippocampus, whereas the previous study^37^ only focussed on anterior regions of hippocampal subfields, and it is possible that posterior sections of subfields may also be hyperactive. Indeed, prior studies reported peak increases in rCBF in CHR-P individuals within the body/tail of the hippocampus^35^, a finding which was replicated in an independent cohort of CHR-P individuals^36^ and in high schizotypy^48^. Additionally, in contrast to the previous study^37^, our study used rCBF which is more tightly coupled to neuronal activity than rCBV^65^, sampled in native vs. common space thereby circumventing normalisation errors^66^, and sampled with individual hippocampal/subfield masks thereby affording higher accuracy given the neuroanatomical diversity among individuals^67^.

In line with our second hypothesis, diazepam significantly reduced hippocampal and subfield rCBF vs. placebo in CHR-P individuals, to the extent that rCBF was no longer significantly different to HC. This finding is consistent with predictions from preclinical studies^29^. In the MAM rodent model, hippocampal hyperactivity is associated with a local reduction of GABAergic PV+ interneurons^24–26^, and increasing GABAergic inhibition by hippocampal infusion of an α5-GABA_A_ PAM normalises hippocampal hyperactivity^29^. Importantly, repeated oral administration of diazepam in peripubertal MAM rats prevents local PV+ interneuron loss^31^ and the emergence of a hyperdopaminergic state at adulthood^30^. A likely mechanism is downregulation of amygdala-hippocampal overdrive, which causes PV+ interneuron loss in the hippocampus and, consequently, hippocampal hyperactivity. This is supported by findings that direct hippocampal infusion of the benzodiazepine midazolam normalises increased dopamine neuron firing in the VTA of adult MAM rats^28^. Our finding of diazepam-induced reductions in rCBF across all hippocampal subfields aligns with the pharmacology of benzodiazepines and GABA_A_ receptor distribution^45,68^. Benzodiazepines are PAMs of the GABA_A_ receptor via the benzodiazepine site, which lies adjacent to α1-3/α5 receptor subunits^69^. Whilst GABA_A_ receptors are expressed on several cell types and sites^70^, most commonly benzodiazepine binding facilitates greater hyperpolarisation of post-synaptic glutamatergic pyramidal cells and reduced pyramidal cell firing^69^. This is expected to result in reduced metabolic requirements and, therefore, reduced rCBF^71^. Although there are slight differences in the levels of α1-3/α5 receptor subunits (and therefore benzodiazepine sites) across hippocampal subfields^68^, α1-3/α5-GABA_A_ receptors are highly expressed across the hippocampus^44,45^, and diazepam has comparably high affinity for them^72^. Additionally, as hippocampal subfields are highly interconnected^69^, it is intuitive that diazepam was associated with a similar magnitude of reduction in rCBF across subfields.

Complementary voxel-wise analyses revealed rCBF reductions across multiple other cortical and subcortical regions in the diazepam condition. The largest reductions were seen in the pre/post central gyrus and inferior frontal regions, which are the areas with the highest benzodiazepine receptor binding sites both in vivo and ex vivo^44^, and receive projections from the hippocampus^15,16^. Large rCBF reductions were also seen in the striatum, ventromedial PFC, and amygdala, which together with the hippocampus compose a cortico-limbic-striatal circuit proposed to be central to the pathophysiology of psychosis^19^. The reductions in rCBF observed in further cortical regions (motor cortex, occipital cortex, thalamus, posterior cingulate cortex) may be related to the ubiquitous binding profile of benzodiazepines. For example, α1-α3 subunits show widespread cortical distribution and have been implicated in benzodiazepine-related side effects such as sedation and addiction^73^. Conversely, α5-GABA_A_ receptors are preferentially expressed in the hippocampus^45,68^, and α5-GABA_A_ agonism of which is not associated with such side effects^73^. Furthermore, there is evidence implicating α5 over α1-3-GABA_A_ receptors in the pathophysiology of psychosis. PET studies in antipsychotic-free patients with schizophrenia using a α5-GABA_A_ selective ligand ([^11^C]-Ro15-4513) showed reduced binding in the hippocampus^74^, and negative correlations with negative symptoms^75^, whilst less specific α1-3,5-GABA_A_ ligands ([^123^I]-iomazenil; [^18^F]-fluoroflumazenil; [^11^C]-flumazenil) found no differences between people with a diagnosis of schizophrenia^76–79^, or CHR-P individuals^80^, and healthy controls. In line with this, evidence from the MAM model shows i) α5 but not α1-3-GABA_A_ receptors are reduced in the subiculum and CA1^81^, ii) overexpression of the α5, but not α1, subunit normalises hippocampal hyperactivity^27^, and iii) an α5-GABA_A_ PAM was able to normalise hippocampal hyperactivity^29^ and attenuate dopamine neuron activity in the VTA to a greater extent that the non-specific benzodiazepine midazolam^28^. Taken together, pharmacological agents with high selectivity for α5-GABA_A_ receptors may be able to regulate hippocampal hyperactivity more specifically in psychosis while potentially avoiding some of the unwanted side effects of less-selective benzodiazepines. While several α5-GABA_A_ PAMs exist^82^, none have yet proceeded to clinical development for psychosis.

Finally, reductions in hippocampal rCBF under diazepam appeared to be smallest in CHR-P participants with higher severity of positive symptoms and poorer social functioning. Current theories propose persistent hyperactivity (i.e., excessive glutamate release) in the hippocampus of CHR-P individuals can lead to atrophic processes of neuropil and PV+ inhibitory interneurons^7^. Therefore, diazepam may not be able to downregulate rCBF as effectively in CHR-P individuals with a more severe clinical profile and who potentially have a greater degree of GABAergic interneuron dysfunction. However, interpretation of this finding is limited by low power for examining correlations between clinical and imaging variables, and differences in timing between clinical and CBF measurements (i.e., 1^st^ and 2^nd^ scan were ∼2 and ∼6 weeks after the assessment visit).

Overall, this novel experimental medicine study presents first-in-human evidence of successful down-regulation of hippocampal hyperactivity by pharmacological modulation of the GABAergic system in CHR-P individuals. We used a robust randomised, double-blind, placebo-controlled, crossover study design, with an adequately powered sample based on prior research determining that n=20 is needed for within-subject pharmacological ASL studies^83^. All participants were antipsychotic-naïve, avoiding the known effects of antipsychotics on rCBF or the GABAergic system^84–86^. We used advanced computational neuroimaging methods to automatically segment the hippocampus into its subfields with high accuracy, using the MAGeT Brain toolbox that outperforms popular competing methods^57^. This high level of accuracy was maintained by sampling ROI rCBF in each participant’s native space and conducting voxel-wise analyses in a study-specific common space. We used ASL, a highly suitable neuroimaging measure (e.g., compared to PET or magnetic resonance spectroscopy [MRS]) for investigating the effects of a GABAergic drug challenge on hippocampal function, given it is fully quantitative, non-invasive, has better spatial resolution and SNR compared to PET^83^, and higher test-retest reliability than GABA concentration measurement using MRS^87^.

This study also has some limitations. Despite segmenting all hippocampal subfields, we could not reliably sample and assess the smaller CA2/3 subfield due to the spatial resolution of ASL. Additionally, unlike the CHR-P group, the HC group were scanned in the absence of a placebo condition. It is possible that placebo administration or broader factors relating to participating in a pharmacological study may have impacted rCBF.

## Conclusions

This study provides first evidence that a single dose of a non-specific GABA-enhancing drug like diazepam can significantly reduce hippocampal and subfield hyperactivity in CHR-P individuals and normalise it to HC levels. Diazepam-associated reductions in rCBF were also observed in other cortico-limbic-striatal regions, supporting further investigation of whether diazepam can modulate the function of this circuit in CHR-P individuals. Furthermore, the results validate the use of ASL and native-space hippocampal and subfield sampling as viable biomarker endpoints for the development of more hippocampal-selective GABA-enhancing compounds for psychosis.

## Data Availability

Data will be made freely available upon publication (10.6084/m9.figshare.24763839), including i) mean rCBF values per subject per ROI per condition, and ii) coding scripts for the MRI preprocessing pipeline (run in Unix/shell) and generation of figures (run in R).

## Acknowledgements

We would like to thank all our participants who took the time to participate in this study, as well as the OASIS clinical team members and the radiographers at the Centre for Neuroimaging Sciences. This is independent research funded by the Wellcome Trust & The Royal Society (grant number 202397/Z/16/Z to GM) and carried out at the National Institute for Health and Care Research (NIHR) Maudsley Biomedical Research Centre (BRC). NRL is supported by an MRC DTP PhD studentship. LAJ is supported by an MRC Clinical Research Training Fellowship (MR/T028084/1). PFP is supported by the European Union funding within the MUR PNRR Extended Partnership initiative on Neuroscience and Neuropharmacology (Project no. PE00000006 CUP H93C22000660006 “MNESYS, A multiscale integrated approach to the study of the nervous system in health and disease”). TJR Is supported by an MRC Clinical Research Training Fellowship (MR/W015943/1). AAG is supported by the National Institute of Mental Health (USPHS MH57440). The views expressed are those of the authors and not necessarily those of the Wellcome Trust & The Royal Society, the NIHR, or the Department of Health and Social Care. For the purpose of open access, the author has applied a Creative Commons Attribution (CC BY) licence to any Accepted Author Manuscript version arising from this submission.

## Author contributions. NRL

Methodology, Software, Formal analysis, Investigation, Data curation, Writing – original draft, Visualization, Project administration; Funding acquisition. **AK:** Investigation, Writing – review & editing, Project administration; **GAD:** Software, Formal analysis, Writing – review & editing; **SK:** Investigation, Writing – review & editing; **PBL:** Investigation, Writing – review & editing; **LJ:** Investigation, Writing – review & editing; **TR:** Investigation, Writing – review & editing; **AD:** Investigation, Writing – review & editing; **MAN:** Investigation, Writing – review & editing; **CC:** Investigation, Writing – review & editing; **TA:** Formal analysis, Writing – review & editing; **FZ:** Software, Resources, Writing – review & editing; **TS:** Resources, Writing – review & editing; **ADM:** Resources, Writing – review & editing; **PFP:** Resources, Writing – review & editing; **AAG:** Writing – review & editing**; SCRW:** Resources, Writing – review & editing; **PM:** Conceptualization, Resources, Writing – review & editing; **AE:** Methodology, Writing – review & editing, Supervision; **MMC:** Software, Methodology, Writing – review & editing, Supervision; **GM:** Conceptualization, Methodology, Writing – review & editing, Supervision, Funding acquisition.

