## Supplementary Material for "Effects of Diazepam on Hippocampal Blood Flow in People at Clinical High Risk for Psychosis"

**SUPPLEMENTARY METHODS ..............................................................................................2**

Clinical and cognitive assessments ...............................................................................2

Scanning visit procedures .............................................................................................2

Further pcASL acquisition details ……............................................................................3

ROI and voxel-wise analysis ..........................................................................................3

Clinical characteristics scores .......................................................................................4

**SUPPLEMENTARY TABLES AND FIGURES .............................................................................5**

Table S1. ROI analyses including different covariates ...................................................5

Figure S1. Visualisation of voxel-wise grey matter rCBF results of CHR-P placebo vs. HC .................................................................................................................................6

Table S2. Statistics of voxel-wise grey matter rCBF results of CHR-P placebo vs. HC ....7

Table S3. Statistics of voxel-wise grey matter rCBF results of CHR-P diazepam vs. placebo .........................................................................................................................8

Table S4. Partial Pearson’s correlations analyses ........................................................10

**SUPPLEMENTAL REFERENCES ...........................................................................................11**

**Supplemental methods**

**Clinical and cognitive assessments**

Several measures were completed at the initial assessment visit to assess clinical symptoms, functioning level, and cognitive ability (table below).

| *Assessment type* | *Assessment domain* | *Assessment name* | *Assessment description* |
| --- | --- | --- | --- |
| Clinical Interview | Clinical Symptoms | Comprehensive Assessment of At Risk Mental State (CAARMS)^1^ | The CAARMS assesses subthreshold psychotic symptoms to determine if an individual meets the criteria for being at CHR-P. |
|  |  | Hamilton Anxiety (HAM-A)^2^ and Depression (HAM-D)^3^ Rating Scales | The HAM-A and HAM-D assess anxiety and depressive symptoms in the last week, respectively. |
|  | Functioning | Global Functioning Social and Role (GF-S/R)^4,5^ | The GF-S/R scales assess social and role functioning individually. |
| Task | Cognition | Wechsler Adult Intelligence Scale – III, Short Version (WAIS-III)^6^ | The shortened WAIS-III allows an individual’s IQ to be estimated using four tasks (Information, Arithmetic, Block Design and Digit Symbol). |
|  |  | Trail Making Test A & B^7^ | This test assesses cognitive flexibility and visuospatial attention. |

**Scanning visit procedures**

If the participant was eligible, they attended two scanning visits at the Centre for Neuroimaging Sciences (CNS), KCL. The participant was informed to avoid recreational substances for 1 week before scanning. They were also asked to avoid alcohol, caffeine, strenuous exercise, operating heavy machinery, or driving for 24 hours before and after the scan, to avoid tobacco-containing products, and to fast for 4 hours before. Procedures for both scanning visits were identical, differing only in whether the participant received 5mg of oral diazepam or placebo (50mg ascorbic acid). A 5mg dose of diazepam was chosen after consultation with neuropsychopharmacologists, indicating that this dose would be sufficient to change brain function without creating substantial subjective effects on the participant/researcher that might have interfered with the maintenance of drug-blinding or led to sedation in the scanner. Randomisation of drug order for the scans (i.e., scan 1 diazepam and scan 2 placebo OR scan 1 placebo and scan 2 diazepam) was achieved using a generalised latin square design. Opaque gel capsules were used for both diazepam and placebo to maintain blinding. Dosing was done 60 minutes prior to the MRI scan to achieve peak plasma levels. MRI scanning lasted a total of 75 minutes and included a battery of sequences. After the scan, given the study doctor's approval that the participant had satisfied ECG, vitals, physical, and neurological examinations, the participant was discharged home. The participant was then scanned again 4 weeks later, receiving the alternative pill.

| **Time** | **Activity** | **Who** | **Location** |
| --- | --- | --- | --- |
| -02:00 | Arrival at CNS | Participant | Clinical Prep Room |
| -01:55 | MRI Safety Form and Request Form | Participant |  |
| -01:45 | VAMS, BSS, PQ-B (Pre-scan) | Participant |  |
| -01:30 | Vitals & Urine Drug and Pregnancy Test | Researcher |  |
| -01:15 | Physical and Neurological Examination | Study Physician |  |
| -01:00 | Dosing with study medication or placebo | Participant |  |
| -00:40 | Take off metallic objects (bra, piercings, etc.) | Participant |  |
| -00:20 | MRI Safety Check | Radiologists |  |
| 00:00 | MRI Scan | Radiologists | CNS 3.0T |
| 01:30 | Out of MRI Scanner | Participant |  |
| 01:35 | Trip to Toilet | Participant |  |
| 01:40 | Snack | Participant |  |
| 01:55 | Physical and Neurological Examination | Study Physician | Clinical Prep Room |
| 02:10 | 12 Lead ECG, Vitals | Researcher |  |
| 02:20 | VAMS, BSS, PQ-B (Post-scan) | Participant |  |
| 02:30 | Discharge if fit to leave | Study Physician |  |
| 02:31 | Discharge instructions | Researcher |  |

**Further pcASL acquisition details**

Arterial blood is labelled using a train of 1000 Hanning-shaped RF pulses with a duration of 500 us and an inter-pulse gap of 1 ms. Following background suppression, arterial blood labelling and a post-labelling delay of 2025 ms, four control-label pair images are acquired using a multi-shot 3D fast spin echo (FSE) spiral readout. Whole-brain imaging is achieved using 56 slices with a slice thickness of 3mm, acquired axially, and stacked on top of each other. Following image reconstruction, this gives a 3D perfusion-weighted difference image with an in-plane resolution of 2x2x3 mm. To compute the CBF map in a standard physiological unit (ml blood/100 g tissue/min) from the difference image, a proton density image is acquired using the same acquisition parameters (TE = 11.088 ms, TR = 4,923 ms, FoV = 240).

**ROI and whole-brain analysis**

Registration of individual ROI masks to individual CBF maps was achieved by calculating the linear transformation matrices to register T1->T2->PD. This multistep registration process was chosen due to the greater similarity in terms of contrast and resolution of T1/T2 and T2/PD compared to T1/PD resulting in superior registration. As the PD and CBF images are aligned in the same space, the concatenation of these transforms was applied to the hippocampal and GM masks to bring them into CBF space in one resampling step in order to avoid multiple resampling errors.

For the generation of the study-specific template, participant-specific averages (using both placebo and diazepam condition structural scans) were first generated before averaging across subjects to generate a study-specific template. The deformation field of each subject’s structural scan to the template was then used to transform each participant’s CBF map from each drug condition to the template and into common space (PD->T2->T1->template) before being smoothed with a 6mm FWHM Gaussian kernel.

**Clinical characteristics scores**

A positive symptom severity score was created by multiplying severity and frequency scores for each positive symptom domain on the CAARMS (P1, P2, P3, P4) and summing these scores together. A negative symptom severity was created using the same formula, but for the negative symptom domains on the CAARMS. A cognitive functioning score reflected by the total time to complete the Trail Making Test Part B. Social and role functioning scores were based on current levels (1-10, with 10 being highest functioning) of functioning on the GF:S/R tool. Anxiety and depression symptom scores were based on the number of scored items on the HAM-A/D, respectively.

**Supplemental Tables and Figures**

**Supplementary Table 1. ROI analyses including different covariates.** Statistical results of supplementary ROI analyses for CHR-P placebo vs. HC and CHR-P diazepam vs. CHR-P placebo including different covariates

| HC vs. CHR-P placebo  adding in covariates IQ, ethnicity, and current cigarette use | Group  F p value | | Group*Hemisphere  F p value | |
| --- | --- | --- | --- | --- |
| Whole hippocampus | 25.827 | **<0.001** | 1.567 | 0.217 |
| CA1 | 27.003 | **<0.001** | 1.067 | 0.307 |
| Subiculum | 25.089 | **<0.001** | 1.225 | 0.274 |
| CA4/DG | 20.934 | **<0.001** | 3.164 | 0.0822 |
| HC vs. CHR-P placebo  adding in ROI volume as covariate | **Group**  **F p value** | | **Group*Hemisphere**  **F p value** | |
| Whole hippocampus | 24.436 | **<0.001** | 1.523 | 0.224 |
| CA1 | 25.432 | **<0.001** | 1.132 | 0.293 |
| Subiculum | 25.078 | **<0.001** | 1.026 | 0.317 |
| CA4/DG | 20.249 | **<0.001** | 3.087 | 0.0861 |
| CHR-P placebo vs. CHR-P diazepam adding in global CBF as covariate | **Drug**  **t value p value** | | **Drug*Hemisphere**  **t value p value** | |
| Whole hippocampus | -2.255 | **0.026995** | 1.040 | 0.302083 |
| CA1 | -2.112 | **0.037958** | 0.971 | 0.335103 |
| Subiculum | -2.414 | **0.0182** | 1.188 | 0.2390 |
| CA4/DG | -2.055 | **0.043319** | 0.929 | 0.356132 |
| CHR-P placebo vs. CHR-P diazepam  adding in covariates global CBF, sex, scan order, and days between scans | **Drug**  **t value p value** | | **Drug*Hemisphere**  **t value p value** | |
| Whole hippocampus | -2.219 | **0.02942** | 1.039 | 0.30238 |
| CA1 | -2.133 | **0.03612** | 0.971 | 0.33491 |
| Subiculum | -2.403 | **0.01872** | 1.188 | 0.23910 |
| CA4/DG | -2.086 | **0.04030** | 0.930 | 0.35586 |

**Supplementary Figure 1. Visualisation of voxel-wise grey matter rCBF results of CHR-P placebo vs. HC**. *T*-statistic map of group (CHR-P placebo vs. HC) effect on rCBF from voxel-wise linear models, controlling for age, sex, and global CBF - thresholded at 5% FDR.

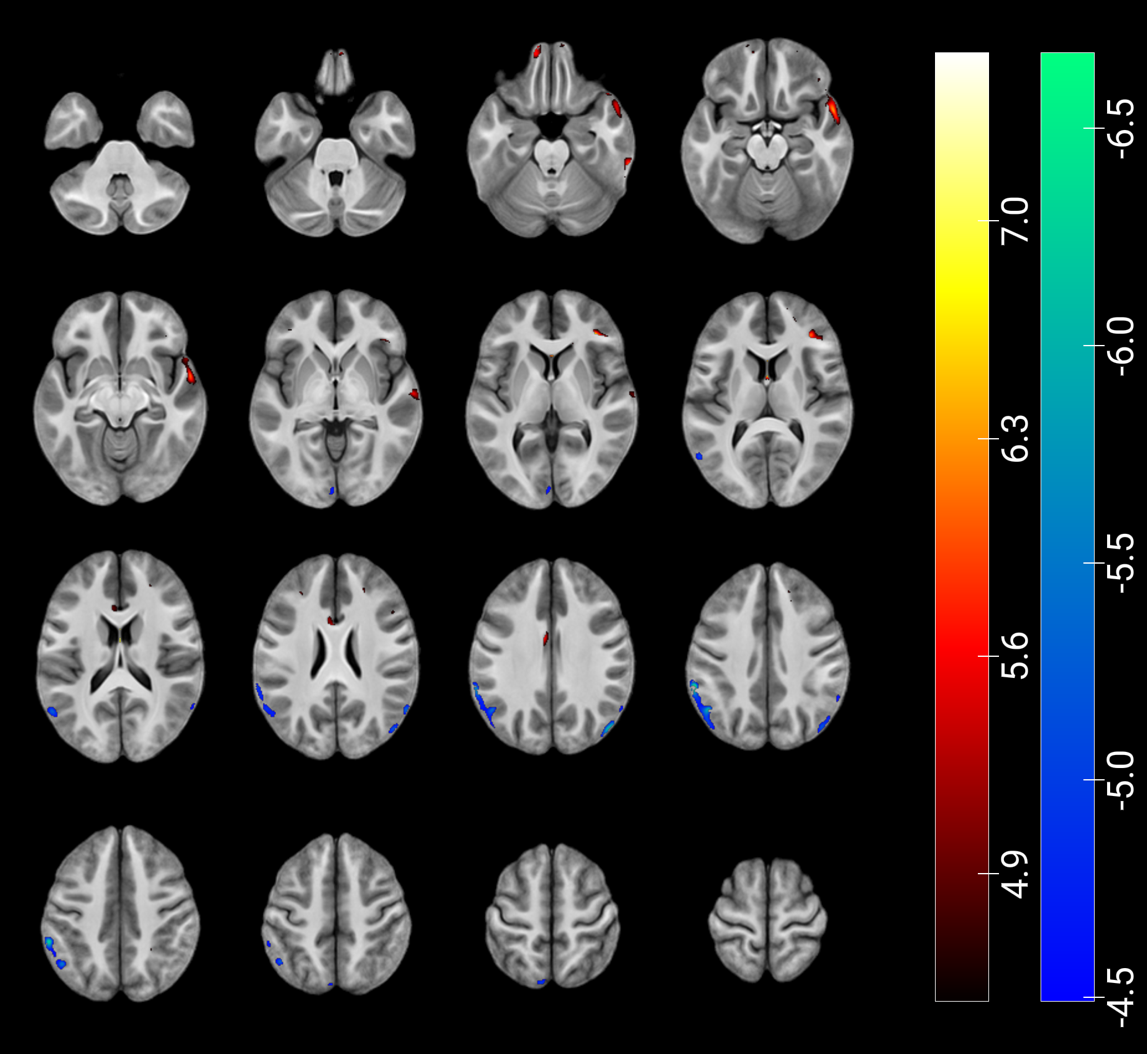

**Supplementary Table 2. Statistics of regions of voxel-wise grey matter rCBF results of CHR-P placebo vs. HC.** Statistical results of voxel-wise effects of group (HC vs. CHR-P placebo) on rCBF thresholded at 5% FDR. N.B. coordinates are in study-specific template space and not MNI coordinates.

| Region | Coordinates | | | t-value | q-value | Number of  peaks |
| --- | --- | --- | --- | --- | --- | --- |
|  | **x** | **y** | **z** |  |  |  |
| CHR-P > HC | | | | | | |
| Right Inferior Frontal Gyrus | 35 | 34 | 7 | 7.036 | 0.009 | 7 |
| Right Inferior Temporal Gyrus | 61 | -34 | -23 | 6.55 | 0.011 | 6 |
| Right Temporal Pole | 53 | 9 | -16 | 6.333 | 0.012 | 3 |
| Left Anterior Orbital Gyrus | -14 | 60 | -19 | 5.969 | 0.012 | 2 |
| Left Medial Orbital Gyrus | -13 | 55 | -22 | 5.836 | 0.012 | 2 |
| Right Gyrus Rectus | 3 | 56 | -26 | 5.582 | 0.014 | 3 |
| Right Superior Temporal Gyrus | 63 | -13 | -1 | 5.533 | 0.015 | 1 |
| Right Lateral Orbital Gyrus | 42 | 32 | -19 | 5.519 | 0.015 | 6 |
| Left Middle Frontal Gyrus | -28 | 32 | 20 | 5.143 | 0.022 | 2 |
| Right Dorsolateral Superior Frontal Gyrus | 16 | 54 | 12 | 5.002 | 0.027 | 7 |
| Right Inferior Frontal Gyrus | 40 | 33 | -11 | 4.991 | 0.027 | 3 |
| Left Dorsolateral Superior Frontal Gyrus | -12 | 54 | -14 | 4.909 | 0.030 | 1 |
| Right Middle Frontal Gyrus | 22 | 35 | 22 | 4.896 | 0.030 | 2 |
| Right Inferior Frontal Gyrus | 47 | 13 | 18 | 4.645 | 0.041 | 1 |
| Left Middle Cingulate | -4 | -4 | 31 | 4.632 | 0.041 | 1 |
| Right Inferior Frontal Gyrus | -37 | 37 | -1 | 4.619 | 0.042 | 1 |
| HC > CHR-P | | | | | | |
| Left Inferior Parietal Gyrus | -58 | -43 | 36 | -6.674 | 0.010 | 3 |
| Left Supramarginal Gyrus | -60 | -46 | 32 | -6.104 | 0.012 | 2 |
| Right Middle Occipital Gyrus | 46 | -76 | 28 | -6.01 | 0.012 | 1 |
| Left Angular Gyrus | -47 | -61 | 36 | -5.979 | 0.012 | 3 |
| Left Middle Temporal Gyrus | -55 | -63 | 13 | -5.412 | 0.017 | 1 |
| Left Precuneus | -9 | -75 | 52 | -4.965 | 0.028 | 1 |
| Left Calcarine Gyrus | -3 | -92 | 0 | -4.931 | 0.029 | 1 |
| Right Middle Temporal Gyrus | 18 | 5 | -41 | -4.717 | 0.037 | 1 |
| Left Inferior Frontal Gyrus | -19 | 22 | 57 | -4.559 | 0.045 | 1 |

**Supplementary Table 3. Statistics of regions of voxel-wise grey matter rCBF results of CHR-P diazepam vs. placebo.** Statistical results of voxel-wise effects of condition (diazepam vs. placebo) on rCBF (without correcting for global CBF) in CHR-P individuals, thresholded at 5% FDR. N.B. coordinates are in study-specific template space and not MNI coordinates.

| Region | Coordinates | | | t-value | q-value | Number of  peaks |
| --- | --- | --- | --- | --- | --- | --- |
|  | **x** | **y** | **z** |  |  |  |
| Right Postcentral Gyrus | 50 | -8 | 30 | -6.68 | 0.003 | 33 |
| Right Superior Temporal Gyrus | 44 | -43 | 14 | -6.442 | 0.003 | 20 |
| Right Inferior Frontal Gyrus (Opercular Part) | 55 | 13 | 8 | -6.428 | 0.003 | 9 |
| Right Dorsolateral Superior Frontal Gyrus | 17 | 13 | 57 | -6.369 | 0.003 | 28 |
| Left Middle Cingulate & Paracingulate Gyri | -3 | -31 | 30 | -6.331 | 0.003 | 10 |
| Right Precentral Gyrus | 36 | -19 | 51 | -6.291 | 0.003 | 34 |
| Right Rolandic Operculum | 49 | 9 | -4 | -6.263 | 0.003 | 10 |
| Left Hippocampus (Head) | -15.5 | 2 | -26 | -6.242 | 0.003 | 4 |
| Left Parahippocampal Gyrus | -16 | 3 | -30 | -6.135 | 0.003 | 3 |
| Right Middle Frontal Gyrus | 32 | 17 | 48 | -6.092 | 0.003 | 29 |
| Right Insula | 42 | -12 | 6 | -5.99 | 0.003 | 13 |
| Right Inferior Frontal Gyrus (Triangular Part) | 51 | 26 | 10 | -5.926 | 0.003 | 11 |
| Left Nucleus Accumbens | -4 | 4 | -12 | -5.811 | 0.003 | 1 |
| Left Putamen | -24 | 9 | -4 | -5.703 | 0.003 | 11 |
| Left Insula | -40 | 13 | -6 | -5.681 | 0.003 | 17 |
| Right Heschl Gyrus | 44 | -13 | 6 | -5.636 | 0.003 | 3 |
| Left Lateral Orbital Gyrus | -40 | 46 | -12 | -5.629 | 0.003 | 4 |
| Left IFG Pars Orbitalis | -41 | 16 | -6 | -5.604 | 0.003 | 4 |
| Right Supramarginal Gyrus | 53 | -45 | 23 | -5.578 | 0.003 | 20 |
| Right IFG Pars Orbitalis | 48 | 26 | -9 | -5.51 | 0.003 | 9 |
| Right Middle Temporal Gyrus | 41 | -53 | 14 | -5.509 | 0.003 | 19 |
| Left Inferior Frontal Gyrus (Opercular Part) | -47 | 12 | 15 | -5.493 | 0.003 | 16 |
| Left Supplementary Motor Area | -2 | 17 | 41 | -5.464 | 0.003 | 16 |
| Left Inferior Frontal Gyrus (Triangular Part) | -53 | 11 | 24 | -5.413 | 0.003 | 17 |
| Right Middle Cingulate & Paracingulate Gyri | 2 | -10 | 36 | -5.306 | 0.003 | 28 |
| Left Postcentral Gyrus | -42 | -12 | 25 | -5.241 | 0.003 | 45 |
| Right Precuneus | 17 | -58 | 16 | -5.234 | 0.003 | 18 |
| Left Temporal Pole: Superior Temporal Gyrus | -27 | 17 | -27 | -5.218 | 0.003 | 6 |
| Right Angular Gyrus | 48 | -53 | 30 | -5.192 | 0.003 | 17 |
| Right Supplementary Motor Area | 3 | 8 | 53 | -5.187 | 0.003 | 13 |
| Right Inferior Temporal Gyrus | 43 | -44 | -16 | -5.021 | 0.003 | 13 |
| Left Lateral Posterior | -6 | -9 | 11 | -4.998 | 0.003 | 2 |
| Right Hippocampus (Head) | 20 | -11 | -20 | -4.911 | 0.003 | 1 |
| Right Hippocampus (Body) | 32 | -18 | -13 | -4.898 | 0.003 | 1 |
| Left Rolandic Operculum | -42 | -14 | 17 | -4.888 | 0.003 | 8 |
| Right Calcarine Fissure | 9 | -88 | 0 | -4.813 | 0.003 | 14 |
| Left Posterior Orbital Gyrus | -28 | 18 | -23 | -4.805 | 0.003 | 1 |
| Right Inferior Parietal Gyrus | 29 | -54 | 40 | -4.796 | 0.003 | 12 |
| Right Parahippocampal Gyrus | 16 | -3 | -30 | -4.764 | 0.003 | 7 |
| Right Lobule VIII Of Cerebellar Hemisphere | 32 | -54 | -52 | -4.706 | 0.003 | 4 |
| Right Hippocampus (Tail) | 14 | -39 | 1 | -4.6 | 0.003 | 4 |
| Left Olfactory Cortex | -18 | 5 | -19 | -4.596 | 0.003 | 1 |
| Left Dorsolateral Superior Frontal Gyrus | -32 | 31 | 34 | -4.583 | 0.003 | 29 |
| Right Nucleus Accumbens | 13 | 14.5 | -11 | -4.578 | 0.003 | 3 |
| Left Precentral Gyrus | -28 | -5 | 43 | -4.569 | 0.003 | 43 |
| Right Medial Superior Frontal Gyrus | 2 | 18 | 41 | -4.547 | 0.003 | 21 |
| Right Putamen | 32 | -19 | -4 | -4.546 | 0.003 | 6 |
| Right Middle Occipital Gyrus | 37 | -74 | 14 | -4.525 | 0.003 | 29 |
| Left Pulvinar Medial | -13 | -23 | 9 | -4.513 | 0.003 | 4 |
| Right Fusiform Gyrus | 37 | -56 | -12 | -4.505 | 0.003 | 6 |
| Left Middle Frontal Gyrus | -26 | -2 | 49 | -4.491 | 0.004 | 17 |
| Left Medial Superior Frontal Gyrus | -2 | 51 | 12 | -4.486 | 0.004 | 25 |
| Left Pallidum | -19 | -5 | -7 | -4.419 | 0.004 | 6 |
| Left Medial Orbital Gyrus | -15 | 7 | -22 | -4.404 | 0.004 | 6 |
| Right Posterior Cingulate Gyrus | 7 | -40 | 6 | -4.373 | 0.004 | 5 |
| Right Lingual Gyrus | 22 | -47 | -6 | -4.357 | 0.004 | 7 |
| Right Caudate Nucleus | 18 | 8 | 17 | -4.354 | 0.004 | 14 |
| Left Fusiform Gyrus | -16 | -39 | -20 | -4.349 | 0.004 | 4 |
| Right Inferior Occipital Gyrus | 33 | -80 | -10 | -4.338 | 0.004 | 7 |
| Left Anterior Orbital Gyrus | -29 | 56 | -9 | -4.337 | 0.004 | 7 |
| Left Anterior Cingulate Cortex (Supracallosal) | -1 | 29 | 29 | -4.291 | 0.004 | 5 |
| Right Pulvinar Anterior | 13 | -28 | -7 | -4.248 | 0.004 | 3 |
| Left Temporal Pole: Middle Temporal Gyrus | -30 | 18 | -37 | -4.244 | 0.004 | 3 |
| Left Caudate Nucleus | -12 | -15 | 22 | -4.224 | 0.004 | 10 |
| Right Anterior Cingulate Cortex (Pregenual) | 3 | 47 | 13 | -4.216 | 0.004 | 4 |
| Right Superior Occipital Gyrus | 24 | -90 | 6 | -4.208 | 0.004 | 25 |
| Left Amygdala | -26 | -4 | -13 | -4.193 | 0.004 | 1 |
| Right Cuneus | 18 | -94 | 6 | -4.172 | 0.004 | 13 |
| Left Posterior Cingulate Gyrus | -6 | -46 | 11 | -4.147 | 0.004 | 1 |
| Left Lobule IV, V Of Cerebellar Hemisphere | -24 | -35 | -28 | -4.145 | 0.004 | 2 |
| Left Parahippocampal Gyrus | -19 | -27 | -20 | -4.139 | 0.004 | 3 |
| Left Precuneus | 0 | -66 | 34 | -4.082 | 0.004 | 17 |
| Right Pulvinar Inferior | 16 | -25 | 6 | -4.024 | 0.004 | 1 |
| Right Pulvinar Medial | 16 | -26 | 3 | -4.009 | 0.005 | 2 |
| Right Posterior Orbital Gyrus | 23 | 22 | -19 | -3.995 | 0.005 | 3 |
| Left Inferior Temporal Gyrus | -33 | -7 | -43 | -3.993 | 0.005 | 11 |
| Right Paracentral Lobule | 13 | -30 | 43 | -3.961 | 0.005 | 2 |
| Left Paracentral Lobule | -18 | -22 | 57 | -3.947 | 0.005 | 16 |
| Left Anterior Cingulate Cortex, Pregenual | -2 | 48 | 7 | -3.922 | 0.005 | 5 |
| Right Temporal Pole: Middle Temporal Gyrus | 50 | 8 | -35 | -3.918 | 0.005 | 1 |
| Left Superior Temporal Gyrus | -59 | -33 | 13 | -3.908 | 0.005 | 10 |
| Right Superior Parietal Gyrus | 27 | -58 | 47 | -3.907 | 0.005 | 12 |
| Left Superior Occipital Gyrus | -10 | -88 | -2 | -3.887 | 0.005 | 15 |
| Left Lingual Gyrus | 0 | -71 | -1 | -3.874 | 0.005 | 6 |
| Left Mediodorsal Medial Magnocellular | -7 | -19 | 1 | -3.861 | 0.005 | 1 |
| Right Crus I Of Cerebellar Hemisphere | 43 | -47 | -37 | -3.773 | 0.005 | 12 |
| Right Gyrus Rectus | 7 | 50 | -18 | -3.76 | 0.005 | 3 |
| Left Calcarine Fissure And Surrounding Cortex | 1 | -81 | -2 | -3.758 | 0.005 | 23 |
| Left Lobule VI Of Cerebellar Hemisphere | -8 | -75 | -22 | -3.749 | 0.005 | 3 |
| Left Middle Temporal Gyrus | -59 | 1 | -17 | -3.749 | 0.005 | 29 |
| Right Crus II Of Cerebellar Hemisphere | 6 | -80 | -34 | -3.749 | 0.005 | 9 |
| Right Medial Orbital Gyrus | 7 | 51 | -20 | -3.728 | 0.006 | 7 |
| Left Crus II Of Cerebellar Hemisphere | -5 | -78 | -31.5 | -3.719 | 0.006 | 9 |
| Left Lobule X Of Cerebellar Hemisphere | -25 | -35 | -38 | -3.714 | 0.006 | 1 |
| Right Amygdala | 27 | -2 | -24 | -3.678 | 0.006 | 1 |
| Left Cuneus | 1 | -76 | 28 | -3.667 | 0.006 | 8 |
| Right Lobule IX Of Cerebellar Hemisphere | 6 | -58 | -47 | -3.654 | 0.006 | 2 |
| Left Inferior Parietal Gyrus | -41 | -33 | 33 | -3.589 | 0.006 | 20 |
| Left Pulvinar Anterior | -13 | -27 | -7 | -3.576 | 0.006 | 3 |
| Right Anterior Cingulate Cortex, Supracallosal | 3 | 24 | 29 | -3.555 | 0.006 | 4 |
| Left Middle Occipital Gyrus | -17 | -95 | -3 | -3.543 | 0.006 | 56 |
| Right Lateral Geniculate | 21 | -22 | -9 | -3.54 | 0.006 | 1 |
| Right Lobule VI Of Cerebellar Hemisphere | 31 | -38 | -35 | -3.522 | 0.006 | 8 |
| Left Superior Frontal Gyrus, Medial Orbital | -2 | 54 | -7 | -3.492 | 0.006 | 5 |
| Left Superior Parietal Gyrus | -17 | -56 | 44 | -3.467 | 0.007 | 20 |
| Right Lenticular Nucleus, Pallidum | 14 | 4 | -2 | -3.464 | 0.007 | 3 |
| Right Lobule X Of Cerebellar Hemisphere | 20 | -40 | -42 | -3.458 | 0.007 | 1 |
| Left Substantia Nigra, Pars Reticulata | -11 | -15 | -16 | -3.444 | 0.007 | 2 |
| Right Superior Frontal Gyrus, Medial Orbital | 1 | 54 | -8 | -3.433 | 0.007 | 6 |
| Left Hippocampus (body) | -24 | -26 | -15 | -3.393 | 0.008 | 1 |
| Right Temporal Pole: Superior Temporal Gyrus | 27 | 7 | -32 | -3.309 | 0.008 | 4 |
| Left Supramarginal Gyrus | -58 | -33 | 24 | -3.292 | 0.008 | 7 |
| Left Gyrus Rectus | -4 | 27 | -23 | -3.272 | 0.009 | 2 |
| Left Raphe Nucleus | 1 | -24 | -17 | -3.267 | 0.009 | 1 |
| Left Lobule IX Of Cerebellar Hemisphere | -11 | -43 | -47 | -3.265 | 0.009 | 1 |
| Left Crus I Of Cerebellar Hemisphere | -22 | -72 | -29 | -3.173 | 0.010 | 7 |
| Right Lobule IV, V Of Cerebellar Hemisphere | 19 | -28 | -26 | -3.061 | 0.012 | 1 |
| Left Angular Gyrus | -33 | -67 | 33 | -3.045 | 0.012 | 12 |
| Right Anterior Orbital Gyrus | 28 | 38 | -13 | -3.008 | 0.012 | 2 |
| Left Inferior Occipital Gyrus | -28 | -85 | -16 | -2.969 | 0.013 | 12 |
| Left Hippocampus (tail) | -22 | -37 | 5 | -2.925 | 0.014 | 1 |
| Left Medial Geniculate | -17 | -23 | -10 | -2.883 | 0.015 | 1 |
| Right Olfactory Cortex | 10 | 10 | -21 | -2.797 | 0.017 | 2 |
| Left Lobule VIII Of Cerebellar Hemisphere | -28 | -59 | -51 | -2.723 | 0.019 | 3 |
| Right Lobule VIIB Of Cerebellar Hemisphere | 40 | -61 | -51 | -2.498 | 0.027 | 1 |

**Supplementary Table 4. Partial Pearson’s correlations analyses.** Partial (controlling for global CBF) Pearson’s correlations for hippocampal rCBF and clinical characteristics

| With mean change in bilateral hippocampal rCBF under diazepam vs. placebo (partial) | r p value | |
| --- | --- | --- |
| CAARMS positive | 0.494 | **0.016** |
| CAARMS negative | -0.293 | 0.208 |
| Cognitive | -0.027 | 0.913 |
| Social functioning | -0.402 | 0.057 |
| Role functioning | -0.217 | 0.319 |
| Anxiety symptoms | -0.192 | 0.405 |
| Depression symptoms | -0.363 | 0.115 |
